# Hyperleukocytosis and outcomes in pediatric B-cell acute lymphoblastic leukemia: A report from the REDIAL Consortium

**DOI:** 10.64898/2026.06.16.26355715

**Authors:** Josiah J. Kim, Austin L. Brown, M. Monica Gramatges, Thanh Hoang, Pagna Sok, Veronica Garcia-Morales, Olga A. Taylor, Van Huynh, Kathleen Ludwig, Laura J. Klesse, Kenneth M. Heym, Timothy Griffin, Rodrigo Erana, John Carlos Bernini, M. Brooke Bernhardt, Philip J. Lupo, Karen R. Rabin, Michael E. Scheurer, Mark Zobeck

## Abstract

Hyperleukocytosis (white blood cell [WBC] count >100 000/µL) at diagnosis is an important prognostic risk factor in pediatric acute lymphoblastic leukemia (ALL), though its significance with contemporary therapy is unclear. We analyzed 1 826 pediatric ALL patients from a multi-institution cohort to determine whether hyperleukocytosis independently predicts outcomes using multivariable Cox proportional hazard modeling. Hyperleukocytosis occurred in 211 patients (12%), with 121 having B-ALL, and showed no prognostic significance in T-ALL patients. In B-ALL, 5-year event-free survival (EFS) was 65% versus 89% for non-hyperleukocytosis patients, and overall survival (OS) was 78% versus 93%. After adjustment for age, cytogenetic risk, central nervous system disease status, and treatment site, hyperleukocytosis remained an independent predictor of end-of-induction minimal residual disease (MRD) positivity (odds ratio 2.53 [95% confidence interval [CI]: 1.71–3.94; p<0.001]), inferior EFS (hazard ratio [HR] 2.44; 95% CI: 1.77–3.38; p<0.001) and inferior OS (HR 2.00; 95% CI: 1.29–3.12; p=0.002). A continuous dose-response relationship was observed between WBC count and these outcomes. Survival associations persisted across all cytogenetic risk categories and MRD strata. Despite risk-adapted therapy with treatment intensification for high-risk features, hyperleukocytosis identifies an aggressive B-ALL phenotype with persistently inferior outcomes, suggesting these patients may benefit from novel therapeutic approaches.

## Introduction

Acute lymphoblastic leukemia (ALL) is the most common pediatric malignancy, with peak incidence occurring in children between the ages of 2 and 5 years.^1^ Advances in risk-adapted therapy have significantly improved survival rates, with 5-year event-free survival (EFS) exceeding 85% in developed countries.^2^ Treatment is stratified based on clinical and genetic risk factors to balance efficacy and toxicity.^3^ High-risk ALL is defined by age at diagnosis (<1 year or >10 years), presenting white blood cell (WBC) count >50 000/µL, specific somatic mutations, and end of induction (EOI) minimal residual disease (MRD) positivity.^4^ These factors guide treatment intensity, though stratification continues to evolve with advancing understanding of the disease’s underlying biology.^5^

Hyperleukocytosis, typically defined as a presenting WBC count >100 000/µL, can lead to life-threatening clinical complications, including tumor lysis syndrome, renal failure, seizures, and respiratory distress.^6^ These acute complications arise from the effects of a high burden of leukemic blasts, but generally resolve with treatment initiation as WBC counts decrease.^7^ The precise mechanisms driving hyperleukocytosis and its relationship with leukemia biology remain incompletely understood.

Current protocols escalate therapy for patients with WBC counts >50 000/µL at diagnosis, typically assigning them to high-risk treatment regimens that include more intensive chemotherapy.^8^ No additional modifications are made for those with even higher WBC counts. A recent study demonstrated that a logarithmic increase in WBC count at diagnosis was predictive of 5-year relapse-free survival for B-ALL.^9^ However, potential biological and clinical mechanisms of this association have not been well explored, and real-world evidence of the association outside the clinical trial setting is lacking.

This study investigates the prognostic impact of hyperleukocytosis in pediatric ALL using real-world data from a multi-institutional patient cohort. We seek to validate the association between hyperleukocytosis and treatment outcomes and explore potential biological and clinical factors that may underlie this relationship. Specifically, we examine whether hyperleukocytosis correlates with other high-risk genetic features, central nervous system (CNS) disease, treatment response metrics such as MRD clearance, and survival outcomes.

## Methods

Data were collected from sites participating in the REducing Disparities in Acute Leukemia (REDIAL) cohort, which is comprised of five pediatric cancer centers in the United States, including Texas Children’s Hospital (TCH; diagnosis years included: 2005 – 2017), Children’s Hospital of Orange County (2010 – 2017), CHRISTUS Children’s Hospital (2006 – 2017), Cook Children’s Hospital (2009 – 2017), and Vannie Cook Children’s Clinic (2005 – 2017). Eligible patients were diagnosed with B- or T-ALL between 1 and 18 years of age.

Institutional Review Boards at each institution reviewed and approved the study protocol.

The demographic variables extracted from the medical records included year of diagnosis, treatment location, age, sex, height, weight, body mass index (BMI; Z-score), and self-reported race and ethnicity. Height and weight were recorded at diagnosis, and BMI Z-scores were computed using the Center for Disease Control age- and sex-specific growth references. Treatment location was analyzed as a binary variable (TCH vs. non-TCH). All patients were treated on or according to Children’s Oncology Group (COG) treatment protocols (Table S1). For B-ALL, the standard-risk protocols included AALL0331 and AALL0932. The high-risk protocols included AALL0232, AALL0622, AALL08P1, AALL07P4, and AALL1131.

For T-ALL, patients were enrolled on AALL0434, AALL1231, or other COG protocols (Table S1). Clinical variables included WBC count at diagnosis (cells/µL), cytogenetic risk categories, CNS disease status, EOI MRD, EFS, and overall survival (OS). Patients were classified as National Cancer Institute (NCI) High-Risk if they met either of the following criteria at diagnosis: age > 10 years or WBC > 50 000/µL. Cytogenetic risk categories were defined as favorable for *ETV6::RUNX1* fusions or double trisomies (DT; trisomies of chromosomes 4 and 10), and unfavorable for Philadelphia chromosome-positive cases (*BCR::ABL1* fusion), intrachromosomal amplification of chromosome 21 (iAMP21), *KMT2A* rearrangements, and hypodiploidy (<44 chromosomes). Cases lacking these changes were categorized as neutral cytogenetics. CNS status was determined via lumbar puncture at diagnosis and defined according to standard COG criteria.^10^ Positive bone marrow EOI MRD was assessed on day 29 and defined as positive when leukemic blasts were detected at ≥0.01% by flow cytometry. Events for the EFS calculation were defined as disease relapse after achieving complete remission, disease progression, or death from any cause.

We performed separate analyses for B-ALL and T-ALL. Descriptive statistics were produced to summarize the demographic and clinical characteristics of the patient population. We used logistic regression to evaluate predictors for the primary outcomes of CNS status and EOI MRD. OS and EFS were analyzed with Cox proportional hazards models. Directed acyclic graphs were developed to determine which covariates to include in multivariable models to estimate the effect of hyperleukocytosis on outcomes. Missing data were imputed using multiple imputation with predictive mean matching.^11,12^ We conducted exploratory pairwise comparisons using Chi-square, Wilcoxon rank-sum, and Log-rank tests for survival outcomes as appropriate. To further examine high-risk patients, we performed subanalyses stratified by WBC category (<50 000/µL, 50 000–100 000/µL, > 100 000/µL at diagnosis), MRD status, and cytogenetic risk category. Additionally, we analyzed log-transformed WBC count as a continuous, nonlinear variable using restricted cubic splines to assess how changes in WBC levels influenced risk. The statistical analyses were performed using R version 4.2.2^13^ (R Foundation for Statistical Computing, Vienna, Austria) with “survival” and “rms” packages.^11,14^

## Results

### Cohort Description

The cohort consisted of 1 826 pediatric ALL patients, of which 1 627 were B-ALL and 199 were T-ALL. There were a total of 211 (12%) presenting with hyperleukocytosis. The median age at diagnosis was 4 years in B-ALL and 9 years in T-ALL, with a male predominance across subtypes. Latino patients comprised the largest racial/ethnic group (58% overall, 67% in B-ALL with hyperleukocytosis). Patients with hyperleukocytosis were treated on or according to COG AALL0232 or AALL1131 (B-ALL, 32% and 59%, respectively) or COG AALL0434 or AALL1231 (T-ALL, 56% and 29%, respectively; Table S1). CNS involvement (CNS2 or CNS3 status) was observed in 19% of patients, and EOI MRD positivity in 25%. Relapse occurred in 238 patients (13%) and death in 159 (9%).

### B-ALL

Among patients with B-ALL, only cytogenetic risk was significantly correlated with hyperleukocytosis at presentation (Table 2). Disease associated with favorable compared to neutral cytogenetic risk group was less likely (odds ratio [OR]: 0.2 [95% confidence interval (CI): 0.14 - 0.42]) to present with hyperleukocytosis. Compared to non-Latino Whites, Asian patients were less likely and non-Hispanic Black patients were more likely to present with hyperleukocytosis, although neither of these associations reached statistical significance. Neither age, BMI, treatment site, nor year of diagnosis were correlated with hyperleukocytosis.

**Table 1:** Demographic, Clinical, and Outcome Characteristics of Pediatric B-ALL and T-ALL Patients Stratified by Hyperleukocytosis Status.

|  | B-ALL (N = 1,627) |  | T-ALL (N = 199) |  |
| --- | --- | --- | --- | --- |
|  | No Hyperleukocytosis (N = 1,506)<sup>1</sup> | Hyperleukocytosis (N = 121)<sup>1</sup> | No Hyperleukocytosis (N = 109)<sup>1</sup> | Hyperleukocytosis (N = 90)<sup>1</sup> |
| **Age** | 4 (3, 9) | 8 (3, 13) | 9 (6, 13) | 9 (4, 13) |
| **Sex** |  |  |  |  |
| F | 681 (45%) | 44 (36%) | 33 (30%) | 27 (30%) |
| M | 825 (55%) | 77 (64%) | 76 (70%) | 63 (70%) |
| **Race/Ethnicity** |  |  |  |  |
| Latino | 868 (58%) | 81 (67%) | 45 (42%) | 36 (40%) |
| Non-Latino White | 441 (30%) | 29 (24%) | 39 (36%) | 29 (33%) |
| Non-Latino Black | 81 (5.5%) | 10 (8.3%) | 11 (10%) | 12 (13%) |
| Non-Latino Asian | 70 (4.7%) | 1 (0.8%) | 8 (7.4%) | 9 (10%) |
| Non-Latino Other | 25 (1.7%) | 0 (0%) | 5 (4.6%) | 3 (3.4%) |
| Missing | 21 | 0 | 1 | 1 |
| **Z Score BMI** | 0.45 (-0.48, 1.38) | 0.85 (-0.07, 1.80) | 0.46 (-0.35, 1.17) | 0.66 (-0.31, 1.53) |
| Missing | 485 | 44 | 43 | 30 |
| **Year of diagnosis** |  |  |  |  |
| 05-09 | 339 (22%) | 21 (17%) | 21 (19%) | 18 (20%) |
| 10-14 | 713 (47%) | 52 (43%) | 54 (50%) | 46 (51%) |
| 15-17 | 464 (31%) | 48 (40%) | 34 (31%) | 26 (29%) |
| **Initial White Blood Cell Count** | 7,590 (3,520, 22,003) | 178,300 (135,140, 280,000) | 22,090 (8,900, 44,400) | 316,415 (178,210, 435,320) |
| **CNS Status** |  |  |  |  |
| 1 | 1,255 (84%) | 68 (57%) | 74 (70%) | 45 (54%) |
| 2 | 207 (14%) | 46 (39%) | 24 (23%) | 27 (32%) |
| 3 | 25 (1.7%) | 5 (4.2%) | 7 (6.7%) | 12 (14%) |
| Missing | 19 | 2 | 4 | 6 |
| **End of Induction MRD** |  |  |  |  |
| <0.01% | 1,113 (77%) | 57 (50%) | 61 (62%) | 47 (55%) |
| ≥0.01% | 330 (23%) | 56 (50%) | 37 (38%) | 38 (45%) |
| Missing | 63 | 9 | 11 | 5 |
| **Relapse** |  |  |  |  |
| No | 1,335 (89%) | 79 (65%) | 97 (89%) | 77 (86%) |
| Yes | 171(11%) | 42 (35%) | 12 (11%) | 13 (14%) |
| **Death** |  |  |  |  |
| No | 1,400 (93%) | 94 (78%) | 95 (87%) | 78 (87%) |
| Yes | 106 (7%) | 27 (22%) | 14 (13%) | 12 (13%) |

<sup>1</sup> Median (IQR); n (%)

**Table 2:** Multivariable Logistic Regression of Demographic and Clinical Predictors of Hyperleukocytosis in Pediatric B-ALL.

| Characteristic | OR<sup>1</sup> | 95% CI<sup>1</sup> | p-value |
| --- | --- | --- | --- |
| Z Score BMI | 1.09 | 0.91, 1.30 | 0.36 |
| Age (years) | 1.04 | 1.00, 1.08 | 0.063 |
| Sex (female) | 0.73 | 0.49, 1.08 | 0.11 |
| Race/Ethnicity |  |  |  |
| Latino | — | — |  |
| Non-Latino Asian | 0.24 | 0.03, 1.82 | 0.17 |
| Non-Latino Black | 1.66 | 0.81, 3.42 | 0.17 |
| Non-Latino White | 0.91 | 0.57, 1.45 | 0.7 |
| Diagnosis year | 1.04 | 0.98, 1.10 | 0.23 |
| Treated at TCH<sup>2</sup> | 0.94 | 0.64, 1.40 | 0.77 |
| Cytogenetic Risk Category |  |  |  |
| Neutral | — | — |  |
| Favorable | 0.24 | 0.14, 0.42 | <0.001 |
| Unfavorable | 1.23 | 0.74, 2.07 | 0.43 |

<sup>1</sup>OR = Odds Ratio, CI = Confidence Interval
<sup>2</sup>TCH = Texas Children's Hospital

Specific cytogenetic lesions were associated with hyperleukocytosis in pairwise analysis.

Patients with hyperleukocytosis were significantly less likely to have the favorable risk cytogenetic markers *ETV6*::*RUNX1* (9.9% with hyperleukocytosis vs. 26% without; Table 3) and DT 4 and 10 (5.8% vs. 23%). Conversely, hyperleukocytosis was more frequently associated with higher-risk markers like *KMT2A* rearrangement (7.4% vs. 1.7%) and *BCR::ABL1* (9.9% vs. 2.4%). Hypodiploidy was not associated with hyperleukocytosis, and iAMP21 was observed significantly more frequently in patients without hyperleukocytosis (0% vs 3.9%).

**Table 3:** Prevalence of Cytogenetic Markers in Patients with and without Hyperleukocytosis.

|  | Variables | Non-Hyperleukocytosis<br>(N = 1,506)<sup>1</sup> | Hyperleukocytosis<br>(N = 121)<sup>1</sup> | p-value<sup>2</sup> |
| --- | --- | --- | --- | --- |
| **Cytogenetic Risk Group** |  |  |  | <0.001 |
|  | Favorable | 701 (47%) | 18 (15%) |  |
|  | Neutral | 143 (9.5%) | 22 (18%) |  |
|  | Unfavorable | 662 (44%) | 81 (67%) |  |
| Favorable | **ETV6::RUNX1** |  |  | <0.001 |
|  | Positive | 396 (26%) | 12 (9.9%) |  |
|  | Negative | 1109 (74%) | 109 (90%) |  |
|  | Unknown | 1 | 0 |  |
|  | **Trisomy 4 & 10** |  |  | <0.001 |
|  | Positive | 342 (23%) | 7 (5.8%) |  |
|  | Negative | 1164 (77%) | 114 (94%) |  |
| Unfavorable | **iAMP21** |  |  | 0.019 |
|  | Positive | 58 (3.9%) | 0 (0%) |  |
|  | Negative | 1447 (96%) | 121 (100%) |  |
|  | Unknown | 1 | 0 |  |
|  | **Hypodiploid** |  |  | >0.9 |
|  | Positive | 24 (1.6%) | 1 (0.8%) |  |
|  | Negative | 1479 (98%) | 120 (99%) |  |
|  | Unknown | 3 | 0 |  |
|  | **KMT2A rearrangement** |  |  | 0.001 |
|  | Positive | 26 (1.7%) | 9 (7.4%) |  |
|  | Negative | 1480 (98%) | 112 (93%) |  |
|  | **Philadelphia Chromosome (BCR::ABL1)** |  |  | <0.001 |
|  | Positive | 36 (2.4%) | 12 (9.9%) |  |
|  | Negative | 1469 (98%) | 109 (90%) |  |
|  | Unknown | 1 | 0 |  |

<sup>1</sup> n (%)
<sup>2</sup> Pearson's Chi-squared test; Fisher's exact test

Three of the four primary outcomes we evaluated were significantly associated with hyperleukocytosis after multivariable adjustment (Table 4). Patients with hyperleukocytosis were more likely to be MRD positive at the end of induction (OR: 2.53 [1.71–3.94]). This group also had poorer EFS (Hazard ratio [HR]: 2.44 [1.77–3.38]) and OS (HR: 2.00 [1.29–3.12]). While a higher proportion of patients with hyperleukocytosis presented with CNS3 status, this association did not reach statistical significance (OR: 1.94 [0.71–5.36]; *p* = 0.20).

**Table 4:** Multivariable Associations of Hyperleukocytosis with CNS Status, EOI Response, and Survival Outcomes.

| Characteristic | OR<sup>1</sup> | 95% CI<sup>1</sup> | p-value | Adjustment Variables |
| --- | --- | --- | --- | --- |
| **CNS3 Status** | 1.94 | 0.71, 5.36 | 0.20 | Z-Score BMI, Age, Sex, Diagnosis Year, Treatment Site, Cytogenetic Risk |
| **EOI MRD** | 2.53 | 1.71, 3.94 | <0.001 | Z-Score BMI, Age, Sex, Diagnosis Year, Treatment Site, Cytogenetic Risk |
|  | HR<sup>1</sup> | 95% CI<sup>1</sup> | p-value | Adjustment Variables |
| **Event Free Survival** | 2.44 | 1.77, 3.38 | <0.001 | Z-Score BMI, Age, Sex, CNS Status, Treatment Site, Diagnosis Year, Cytogenetic Risk |
| **Overall Survival** | 2.00 | 1.29, 3.12 | 0.0021 | Z-Score BMI, Age, Sex, CNS Status, Treatment Site, Diagnosis Year, Cytogenetic Risk |

<sup>1</sup>OR, Odds Ratio. HR, Hazard Ratio, CI = Confidence Interval

When hyperleukocytosis was analyzed across the categories ≤ 50 000/µL, 50 000 – 100 000/µL, and ≥ 100 000/µL, higher WBC category was associated with decreased EFS and OS on the log-rank test (Figure 1A,B). The 5-year EFS was 86% for the 1 360 patients with <50 000/µL WBC at diagnosis, 75.7% for 146 patients with 50 000–100 000/µL, and 53.3% for 121 patients with >100 000/µL. Similarly, the 5-year OS by increasing WBC category was 93.7%, 85.8%, and 72.8%, respectively. After further stratification by MRD status (Figure 1C–F), patients who were MRD positive at the end of induction demonstrated a dose-dependent decrease in EFS (log-rank *p*<0.0001) and OS (*p*<0.0001). However, among MRD-negative patients, only WBC counts >100 000/µL remained associated with inferior EFS (*p*<0.0001) and OS (*p*<0.0001). A similar pattern persisted among the 580 patients who met NCI High-Risk criteria, where only WBC >100 000/µL demonstrated lower EFS (p<0.0001) and OS (p<0.024) compared to the other groups (Figure 1G,H).

**Figure 1.**
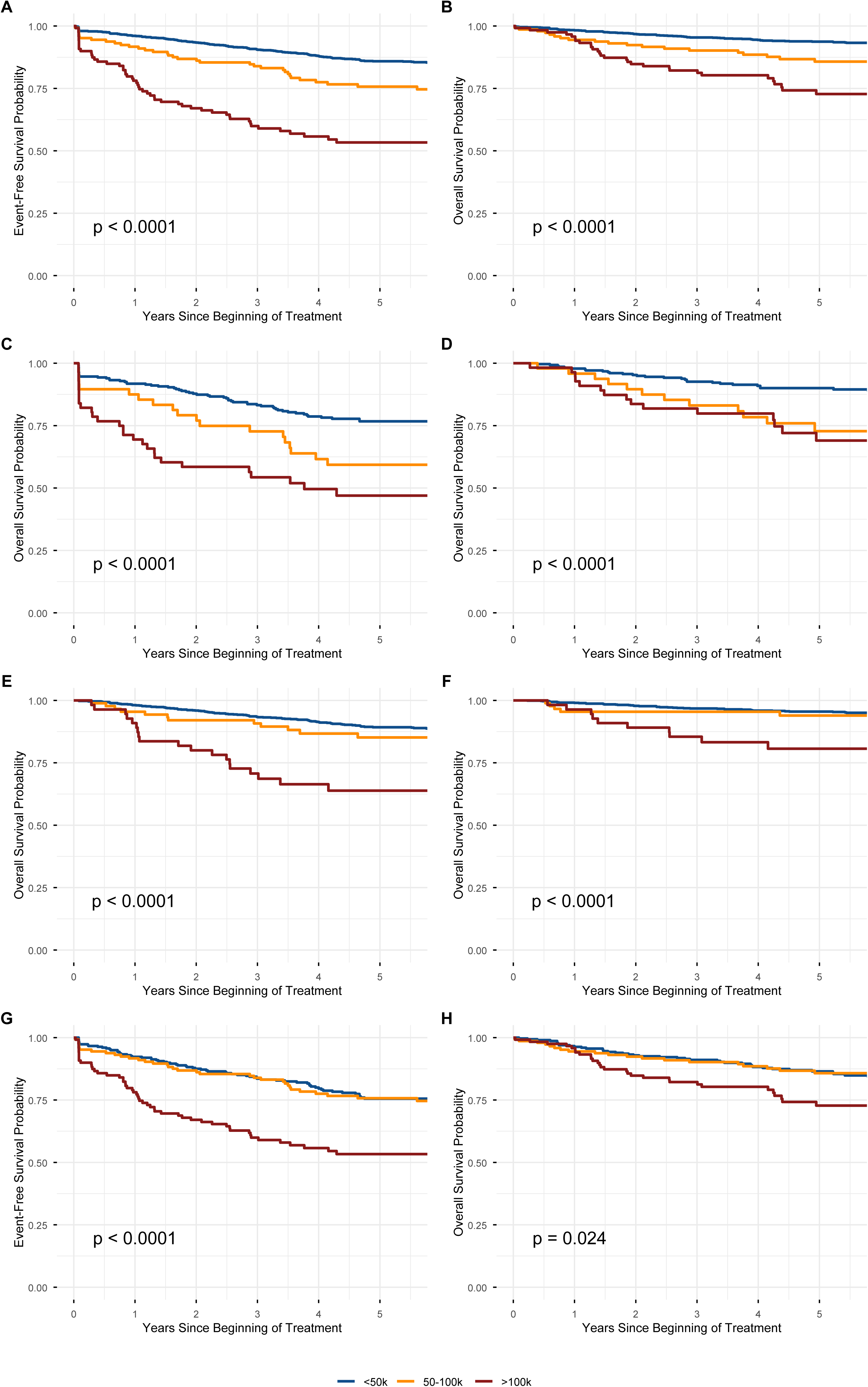
Event-Free and Overall Survival by white blood cell count category for the entire cohort (A, B), EOI MRD-positive patients (C, D), MRD-negative patients (E, F), and National Cancer Institute high-risk patients (WBC >50 000/µL or age ≥10 years; G, H). WBC categories: <50 000/µL (n=1 360), 50 000–100 000/µL (n=146), and >100 000/µL (n=121).

Across all cytogenetic risk categories, patients with hyperleukocytosis had significantly lower EFS (Figure 2). Patients with favorable or neutral cytogenetics with hyperleukocytosis had worse OS vs. non-hyperleukocytosis, whereas patients with unfavorable risk cytogenetics had a similar OS regardless of hyperleukocytosis (Figure 2). Finally, after multivariable adjustment by age and covariate terms for each individual cytogenetic lesion listed in Table 4, hyperleukocytosis remained significant for lower EFS (HR: 2.44 [1.77–3.38]; *p*<0.001) and OS (HR: 2.00 [1.29–3.12]; *p*=0.002).

**Figure 2.**
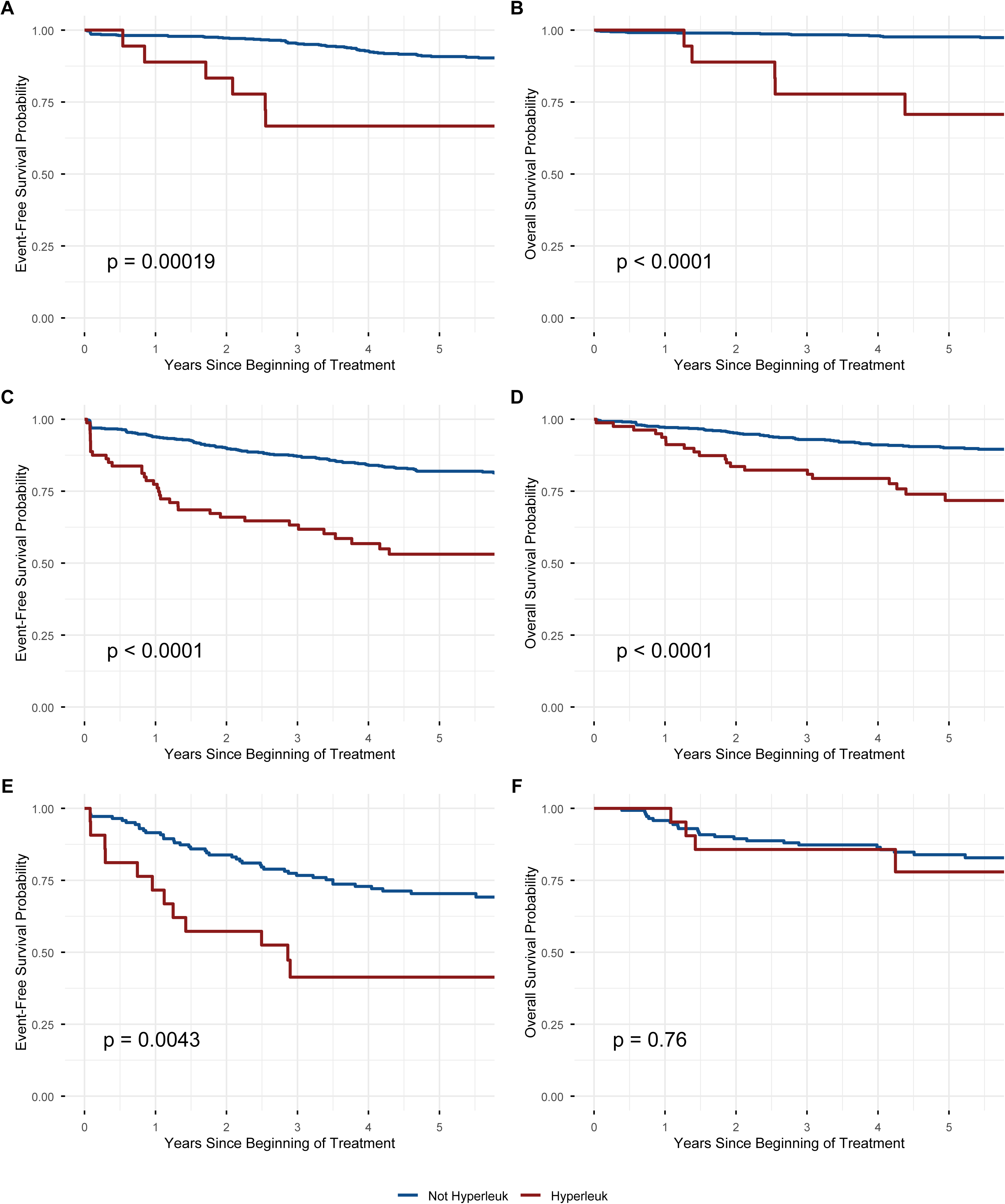
Event-Free and Overall Survival by favorable (A, B), neutral (C, D), and unfavorable (E, F) cytogenetic risk, comparing patients with and without hyperleukocytosis.

Analyzing log_2_ WBC counts at diagnosis revealed a positive continuous dose-response relationship between increasing WBC at diagnosis and risk of adverse outcomes (Figure 3). The log-linear associations were statistically significant for MRD positivity (*p*<0.001), 5-year EFS (*p*<0.001), and 5-year OS (*p*<0.001). The association for CNS3 status did not reach statistical significance (*p*=0.20). MRD positivity was the only outcome with a significant nonlinear association (*p*=0.006), with risk that increased sharply at approximately 10 000 WBC/µL (Figure 3).

**Figure 3.**
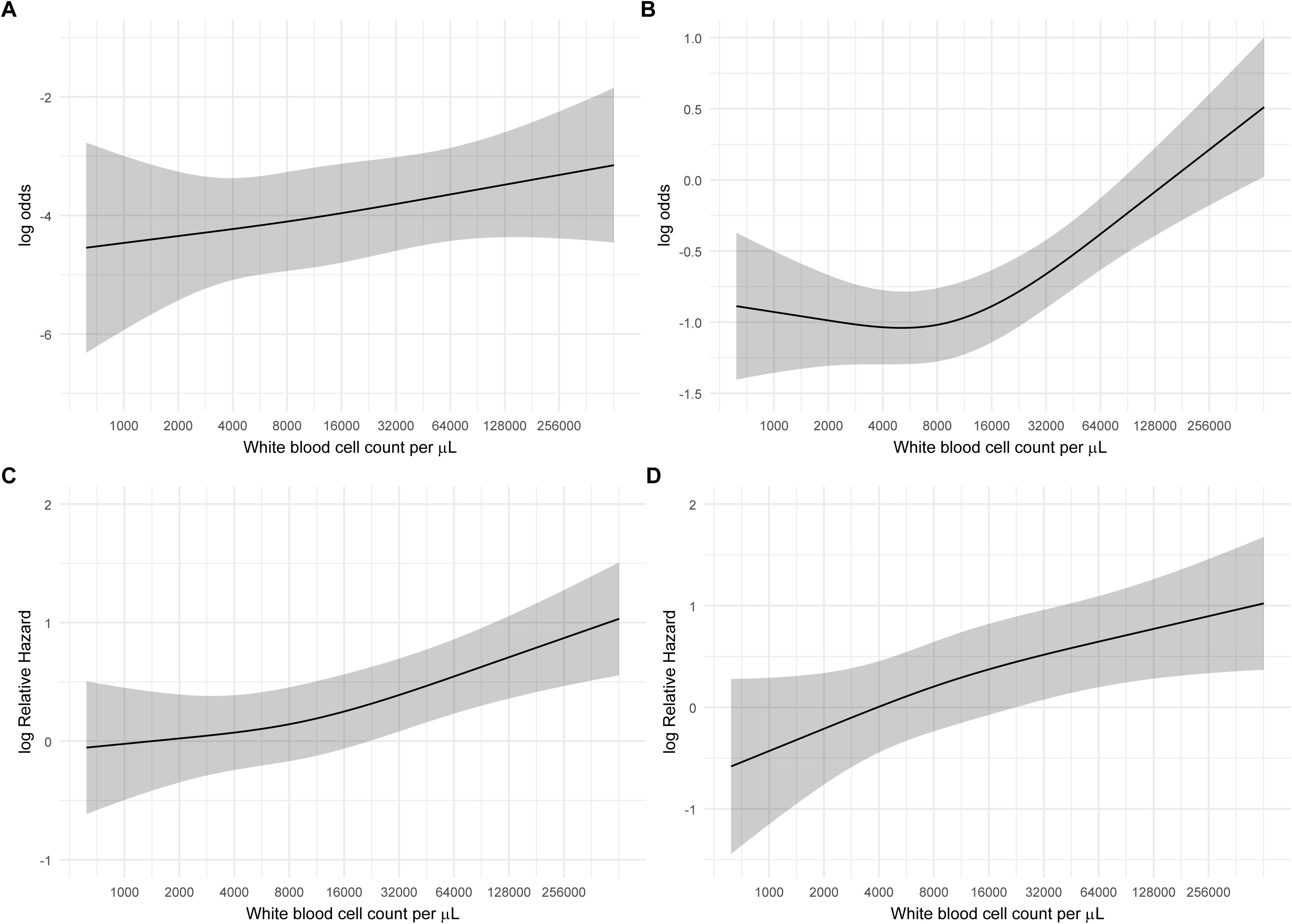
Odds/Hazard Ratios for A) CNS-3 status, B) Positive EOI MRD, C) EFS, and D) OS by log2(WBC) modeled nonlinearly using restricted cubic splines (3 knots). Shaded regions represent 95% confidence intervals.

### T-ALL

No significant risk factors for the hyperleukocytosis at diagnosis for T-ALL were identified (Table S2). On multivariable analysis, hyperleukocytosis was not significantly associated with adverse outcomes in patients with T-ALL (Table S3). Hyperleukocytosis showed a trend toward higher odds of CNS3 status (OR: 2.44; [0.89–6.68]; p=0.08), but no clear association with EOI MRD positivity (OR: 1.3 [0.71–2.38]; p=0.4). Hyperleukocytosis was not independently associated with EFS (HR: 0.52; 95% CI: 0.19–1.45; p=0.21) or OS (HR: 1.0; 95% CI: 0.29–3.44; p=0.99; Figure S1). Three of four patients (75%) positive for *BCR::ABL1* presented with hyperleukocytosis compared to 87 of 195 (45%) *BCR::ABL1-*negative patients (Fisher’s exact *p*=0.3). Three of six patients (50%) with *KMT2A* rearrangement presented with hyperleukocytosis compared to 87 of 193 (45%) without *KMT2A* rearrangement (Fisher’s exact p>0.9).

## Discussion

In a large, multicenter cohort of pediatric patients with ALL, hyperleukocytosis at diagnosis was strongly associated with worse EFS and OS in pediatric patients with B-ALL, independent of other prognostic factors. Despite intensification of induction therapy for patients with high-risk features at diagnosis, hyperleukocytosis at diagnosis remained significantly associated with positive EOI MRD. Moreover, despite intensification of post-induction therapy for patients with high-risk features at diagnosis as well as for patients with unfavorable cytogenetic features and/or EOI MRD positivity, hyperleukocytosis was predictive of EFS and OS in the overall B-ALL cohort and remained predictive across subgroups stratified by WBC category, MRD status, and cytogenetic risk group. Taken together, these observations suggest that hyperleukocytosis reflects an aggressive disease phenotype for B-ALL that increases the risk of treatment failure despite therapy intensification for high-risk disease.

Evidence from other studies supports this conclusion. Early investigations in the 1980s and 1990s among patients with ALL identified a threshold of 200 000 cells/µL as prognostic for poorer OS.^15–19^ Hastings, et al., evaluated patients treated from 1996–2002 on the Children’s Cancer Group (COG) CCG-1961 protocol^20^ and demonstrated a WBC threshold of 200 000/µL as prognostic for B-ALL patients (5-year EFS 47.9% vs 73.0% for non-markedly elevated), whereas no difference in outcomes by WBC was observed for T-ALL (72.0% vs 73.7%) after post-induction treatment intensification. More recently, DelRocco, et al., reaffirmed the prognostic significance of WBC count at diagnosis in the modern era in data from four COG trials (AALL0331, AALL0232, AALL0932, and AALL1131; 2004–2019) as a continuous variable in the prognosis prediction model.^9^ In a similar analysis, Enshaei, et al., derived and externally validated a continuous prognostic index (PIUKALL) that integrates presenting WBC with pretreatment cytogenetics and end-of-induction MRD.^21^ These findings support the clinical relevance of hyperleukocytosis as a marker of aggressive disease biology across different treatment eras and approaches to risk stratification.

This study advances our understanding of the prognostic importance of hyperleukocytosis in several key ways. We provided evidence for a continuous and non-linear dose-dependent relationship between WBC count and adverse outcomes, with risk increasing progressively rather than at discrete thresholds. The risk of positive MRD increased nonlinearly with WBC count, with a steep rise starting around 10 000 cells/µL, indicating an accelerating risk of induction failure as WBC count increases. These findings suggest that a traditional WBC threshold for treatment intensification of 50 000 cells/µL at diagnosis may fail to consider a population at increased risk for treatment failure. Alternative therapy intensification strategies to account for these risk dynamics should be explored.

We found that risk for MRD positivity contributed to, but did not completely explain, the association between hyperleukocytosis and reduced survival. Hyperleukocytosis was associated with an increased risk of EOI MRD positivity, a known prognostic factor for treatment failure and poor survival.^22–24^ White blood cell counts between 50 000 – 100 000/µL and > 100 000/µL were predictive of lower OS among MRD-positive patients, and WBC > 100 000/µL had worse outcomes among MRD-negative patients. This suggests that hyperleukocytosis may represent both a risk for inadequate cytoreduction during induction therapy as well as a marker of resistant disease even among those who have adequate response to initial treatment.

Our analysis demonstrated associations between hyperleukocytosis and specific cytogenetic lesions. Patients with hyperleukocytosis were significantly less likely to harbor favorable genetic markers such as *ETV6::RUNX1* fusions and double trisomies, while demonstrating increased prevalence of high-risk alterations such as *KMT2A* rearrangements and *BCR::ABL1* fusions. Despite these associations, hyperleukocytosis remained an independent prognostic factor even after adjusting for cytogenetic risk categories. One possible explanation for this may be the presence of additional genetic drivers not captured by current standard cytogenetic testing. Of note, other *BCR::ABL1*-like alterations were not systematically analyzed in our cohort,^25^ and neither were novel groups of driver genes in pediatric ALL, such as *IKZF1*plus, *PAX5*alt, *MEF2D* rearranged, and *ETV6::RUNX1*-like ALL.^26–28^ To better understand the association between hyperleukocytosis and poor outcomes, further work is needed to evaluate the hypothesis that unidentified driver genes of poor outcomes are also related to hyperleukocytosis.

An alternative explanation for the association between hyperleukocytosis and poor outcomes may lie in the pathophysiological mechanisms associated with dense concentrations of leukemia cells. Leukemia blasts disrupt the bone marrow microenvironment through various mechanisms, including complex interactions with mesenchymal stem cells and hypoxia-induced endothelial cell disruption.^29,30^ In AML, researchers have theorized that hyperleukocytosis-associated treatment resistance may result from enhanced leukemic cell-stromal interactions mediated by cell surface receptors, such as CD43, CD44, CXCR4, and VCAM-1, which disrupt normal bone marrow architecture and create chemotherapy-resistant niches.^33^ Because these markers are similarly expressed in lymphoid blasts,^34–37^ analogous mechanisms may underlie the effects of hyperleukocytosis in B-ALL and warrant further investigation.

Our study has several limitations. No patients in this study were treated with frontline blinatumomab, which improves survival among pediatric patients with standard-risk B-ALL and adults with MRD-negative ALL.^38,39^ Thus, the prognostic significance of hyperleukocytosis for patients who receive blinatumomab remains unknown. As a retrospective analysis, our study is subject to potential biases and unobserved confounding. Some patients with hyperleukocytosis in critical condition at presentation were assigned a CNS status without obtaining pre-treatment cerebrospinal fluid, potentially leading to misclassification. Additionally, limited sample sizes for certain disease subtypes, such as *CRLF2* rearrangements and infant ALL, restrict generalization of the findings in these groups.

In conclusion, hyperleukocytosis is a strong independent predictor of worse outcomes in pediatric B-ALL. The association persists across MRD strata and cytogenetic risk groups, suggesting that hyperleukocytosis reflects an aggressive disease phenotype that remains incompletely understood. No significant associations were observed in T-ALL, suggesting distinct biological mechanisms between leukemia subtypes, though limited sample size may also contribute to this finding. These findings raise the question of what underlying genetic and pathophysiological mechanisms drive effects associated with hyperleukocytosis. Additional studies are needed to validate this association on contemporary immunotherapy-containing protocols and to identify a functional basis for targeted strategies to improve outcomes.

## Acknowledgements

This work was supported in part by the Cancer Prevention & Research Institute of Texas (CPRIT; Core Facility Support Awards RP210064 and RP160771 to M.E.S./T.T.H.), the National Cancer Institute (P20CA262733 and U54CA302464 to K.R.R. and P.J.L., K12CA090433 to M.Z.), and St. Baldrick’s Foundation Consortium Research Grant 522277 with support from the Micaela’s Army Foundation (P.J.L. and K.R.R.). K.R.R. was supported in part by the Lynch family and is the Deborah and Arthur Ablin Chair in Pediatric Molecular Oncology at the UCSF School of Medicine.

## Author Contributions

M.Z. and J.J.K. conceived and designed the study. M.Z., A.L.B., M.M.G., T.H., P.S., V.G-M., O.A.T., V.H., K.L., L.J.K., K.M.H., T.G., R.E., J.C.B., M.B.B., P.J.L., K.R.R., and M.E.S. contributed to data acquisition. J.J.K. and M.Z. performed data analysis and statistical modeling.

J.J.K. and M.Z. drafted the manuscript. All authors reviewed, revised, and approved the final manuscript.

## Data Availability Statement

The data that support the findings of this study contain protected health information from pediatric patients across multiple institutions and are not publicly available due to participant confidentiality and Institutional Review Board restrictions. De-identified summary data that support the findings of this study are available from the corresponding author upon reasonable request, subject to institutional and data-sharing agreement approvals.

---

**Figure S1.** Event-Free Survival (A) and Overall Survival (B) for patients with T-ALL, comparing those with (n=90) and without (n=109) hyperleukocytosis at diagnosis.

## References

1. Hunger SP, Mullighan CG. Acute Lymphoblastic Leukemia in Children. N Engl J Med. 2015 Oct 15;373(16):1541–52.

2. Pui CH, Yang JJ, Hunger SP, Pieters R, Schrappe M, Biondi A, et al. Childhood Acute Lymphoblastic Leukemia: Progress Through Collaboration. J Clin Oncol Off J Am Soc Clin Oncol. 2015 Sept 20;33(27):2938–48.

3. Inaba H, Greaves M, Mullighan CG. Acute lymphoblastic leukaemia. Lancet Lond Engl. 2013 June 1;381(9881):1943–55.

4. Borowitz MJ, Devidas M, Hunger SP, Bowman WP, Carroll AJ, Carroll WL, et al. Clinical significance of minimal residual disease in childhood acute lymphoblastic leukemia and its relationship to other prognostic factors: a Children’s Oncology Group study. Blood. 2008 June 15;111(12):5477–85.

5. Mullighan CG. Genomic Characterization of Childhood Acute Lymphoblastic Leukemia. Semin Hematol. 2013 Oct 1;50(4):314–24.

6. Howard SC, Jones DP, Pui CH. The Tumor Lysis Syndrome. N Engl J Med. 2011 May 12;364(19):1844–54.

7. Pui CH, Robison LL, Look AT. Acute lymphoblastic leukaemia. Lancet Lond Engl. 2008 Mar 22;371(9617):1030–43.

8. Pui CH, Pei D, Raimondi SC, Coustan-Smith E, Jeha S, Cheng C, et al. Clinical impact of minimal residual disease in children with different subtypes of acute lymphoblastic leukemia treated with Response-Adapted therapy. Leukemia. 2017 Feb;31(2):333–9.

9. DelRocco NJ, Loh ML, Borowitz MJ, Gupta S, Rabin KR, Zweidler-McKay P, et al. Enhanced Risk Stratification for Children and Young Adults with B-Cell Acute Lymphoblastic Leukemia: A Children’s Oncology Group Report. Leukemia. 2024 Apr;38(4):720–8.

10. Winick N, Devidas M, Chen S, Maloney K, Larsen E, Mattano L, et al. Impact of Initial CSF Findings on Outcome Among Patients With National Cancer Institute Standard- and High-Risk B-Cell Acute Lymphoblastic Leukemia: A Report From the Children’s Oncology Group. J Clin Oncol Off J Am Soc Clin Oncol. 2017 Aug 1;35(22):2527–34.

11. Jr FEH. rms: Regression Modeling Strategies [Internet]. 2024 [cited 2024 Nov 10]. Available from: https://cran.r-project.org/web/packages/rms/index.html

12. Jr FEH, functions) CD (contributed several functions and maintains latex. Hmisc: Harrell Miscellaneous [Internet]. 2024 [cited 2024 Nov 10]. Available from: https://cran.r-project.org/web/packages/Hmisc/

13. R Core Team. R: A Language and Environment for Statistical Computing [Internet]. Vienna, Austria: R Foundation for Statistical Computing; 2024. Available from: https://www.R-project.org/

14. Therneau TM, Grambsch PM. Modeling Survival Data: Extending the Cox Model. New York: Springer; 2000.

15. Eguiguren J, Schell M, Crist W, Kunkel K, Rivera G. Complications and outcome in childhood acute lymphoblastic leukemia with hyperleukocytosis. Blood. 1992 Feb 15;79(4):871–5.

16. Nachman J, Sather HN, Gaynon PS, Lukens JN, Wolff L, Trigg ME. Augmented Berlin-Frankfurt-Munster therapy abrogates the adverse prognostic significance of slow early response to induction chemotherapy for children and adolescents with acute lymphoblastic leukemia and unfavorable presenting features: a report from the Children’s Cancer Group. J Clin Oncol. 1997 June;15(6):2222–30.

17. Steinherz PG, Gaynon PS, Breneman JC, Cherlow JM, Grossman NJ, Kersey JH, et al. Treatment of patients with acute lymphoblastic leukemia with bulky extramedullary disease and T-cell phenotype or other poor prognostic features. Cancer. 1998;82(3):600–12.

18. Saarinen-Pihkala UM, Gustafsson G, Carlsen N, Flaegstad T, Forestier E, Glomstein A, et al. Outcome of children with high-risk acute lymphoblastic leukemia (HR-ALL): Nordic results on an intensive regimen with restricted central nervous system irradiation. Pediatr Blood Cancer. 2004;42(1):8–23.

19. Schultz KR, Pullen DJ, Sather HN, Shuster JJ, Devidas M, Borowitz MJ, et al. Risk- and response-based classification of childhood B-precursor acute lymphoblastic leukemia: a combined analysis of prognostic markers from the Pediatric Oncology Group (POG) and Children’s Cancer Group (CCG). Blood. 2006 Sept 26;109(3):926–35.

20. Hastings C, Gaynon PS, Nachman JB, Sather HN, Lu X, Devidas M, et al. Increased post-induction intensification improves outcome in children and adolescents with a markedly elevated white blood cell count (≥200 × 109/l) with T cell acute lymphoblastic leukaemia but not B cell disease: a report from the Children’s Oncology Group. Br J Haematol. 2015;168(4):533–46.

21. Enshaei A, O’Connor D, Bartram J, Hancock J, Harrison CJ, Hough R, et al. A validated novel continuous prognostic index to deliver stratified medicine in pediatric acute lymphoblastic leukemia. Blood. 2020 Apr 23;135(17):1438–46.

22. Panzer-Grümayer ER, Schneider M, Panzer S, Fasching K, Gadner H. Rapid molecular response during early induction chemotherapy predicts a good outcome in childhood acute lymphoblastic leukemia. Blood. 2000 Feb 1;95(3):790–4.

23. Borowitz MJ, Wood BL, Devidas M, Loh ML, Raetz EA, Salzer WL, et al. Prognostic significance of minimal residual disease in high risk B-ALL: a report from Children’s Oncology Group study AALL0232. Blood. 2015 Aug 20;126(8):964–71.

24. Berry DA, Zhou S, Higley H, Mukundan L, Fu S, Reaman GH, et al. Association of Minimal Residual Disease With Clinical Outcome in Pediatric and Adult Acute Lymphoblastic Leukemia: A Meta-analysis. JAMA Oncol. 2017 July 13;3(7):e170580.

25. Tasian SK, Loh ML, Hunger SP. Philadelphia chromosome–like acute lymphoblastic leukemia. Blood. 2017 Nov 9;130(19):2064–72.

26. Jeha S, Choi J, Roberts KG, Pei D, Coustan-Smith E, Inaba H, et al. Clinical Significance of Novel Subtypes of Acute Lymphoblastic Leukemia in the Context of Minimal Residual Disease–Directed Therapy. Blood Cancer Discov. 2021 July 1;2(4):326–37.

27. Brady SW, Roberts KG, Gu Z, Shi L, Pounds S, Pei D, et al. The genomic landscape of pediatric acute lymphoblastic leukemia. Nat Genet. 2022 Sept;54(9):1376–89.

28. Stanulla M, Dagdan E, Zaliova M, Möricke A, Palmi C, Cazzaniga G, et al. IKZF1plus Defines a New Minimal Residual Disease-Dependent Very-Poor Prognostic Profile in Pediatric B-Cell Precursor Acute Lymphoblastic Leukemia. J Clin Oncol Off J Am Soc Clin Oncol. 2018 Apr 20;36(12):1240–9.

29. Witkowski MT, Kousteni S, Aifantis I. Mapping and targeting of the leukemic microenvironment. J Exp Med. 2019 Dec 24;217(2):e20190589.

30. Ciciarello M, Corradi G, Forte D, Cavo M, Curti A. Emerging Bone Marrow Microenvironment-Driven Mechanisms of Drug Resistance in Acute Myeloid Leukemia: Tangle or Chance? Cancers. 2021 Oct 22;13(21):5319.

31. Spertini C, Baïsse B, Bellone M, Gikic M, Smirnova T, Spertini O. Acute Myeloid and Lymphoblastic Leukemia Cell Interactions with Endothelial Selectins: Critical Role of PSGL-1, CD44 and CD43. Cancers. 2019 Sept;11(9):1253.

32. Fodil S, Arnaud M, Vaganay C, Puissant A, Lengline E, Mooney N, et al. Endothelial cells: major players in acute myeloid leukaemia. Blood Rev. 2022 July 1;54:100932.

33. Bewersdorf JP, Zeidan AM. Hyperleukocytosis and Leukostasis in Acute Myeloid Leukemia: Can a Better Understanding of the Underlying Molecular Pathophysiology Lead to Novel Treatments? Cells. 2020 Oct;9(10):2310.

34. Jacamo R, Chen Y, Wang Z, Ma W, Zhang M, Spaeth EL, et al. Reciprocal leukemia-stroma VCAM-1/VLA-4-dependent activation of NF-κB mediates chemoresistance. Blood. 2014 Apr 24;123(17):2691–702.

35. Giorgiutti S, Rottura J, Korganow AS, Gies V. CXCR4: from B-cell development to B cell–mediated diseases. Life Sci Alliance. 2024 Mar 22;7(6):e202302465.

36. Corcione A, Arduino N, Ferretti E, Pistorio A, Spinelli M, Ottonello L, et al. Chemokine receptor expression and function in childhood acute lymphoblastic leukemia of B-lineage. Leuk Res. 2006 Apr 1;30(4):365–72.

37. Abobakr A, Osman RA, Kamal MAM, Abdelhameed S, Ismail H, Kamel MM, et al. Clinical and prognostic significance of CD27 and CD44 expression patterns in Egyptian pediatric patients with B-precursor acute lymphoblastic leukemia. Hematol Transfus Cell Ther. 2024 Dec 1;46:S27–35.

38. Litzow MR, Sun Z, Mattison RJ, Paietta EM, Roberts KG, Zhang Y, et al. Blinatumomab for MRD-Negative Acute Lymphoblastic Leukemia in Adults. N Engl J Med. 2024 July 25;391(4):320–33.

39. Gupta S, Rau RE, Kairalla JA, Rabin KR, Wang C, Angiolillo AL, et al. Blinatumomab in Standard-Risk B-Cell Acute Lymphoblastic Leukemia in Children. N Engl J Med. 2025 Feb 27;392(9):875–91.

