## Supplemental Figures for "Hyperleukocytosis and outcomes in pediatric B-cell acute lymphoblastic leukemia: A report from the REDIAL Consortium"

**Supplement**

**Table S1.** Induction protocols for patients in the cohort.

| **Protocol** | **Non-Hyperleukocytosis** | **Hyperleukocytosis** |
| --- | --- | --- |
| **B ALL** | | |
| AALL0232 | 177 (12%) | 39 (32%) |
| AALL0331 | 329 (22%) | 0 (0%) |
| AALL0622 | 3 (0.2%) | 0 (0%) |
| AALL07P4 | 9 (0.6%) | 7 (5.8%) |
| AALL08P1 | 11 (0.7%) | 4 (3.3%) |
| AALL0932 | 672 (45%) | 0 (0%) |
| AALL1131 | 305 (20%) | 71 (59%) |
| **T ALL** | | |
| AALL0434 | 66 (61%) | 50 (56%) |
| AALL1231 | 30 (28%) | 26 (29%) |
| Other | 13 (12%) | 14 (16%) |

**Table S2: Demographic and Clinical Predictors of T-ALL Hyperleukocytosis**

| **Variable** | **OR***^1^* | **95% CI***^1^* | **p-value** |
| --- | --- | --- | --- |
| **Z Score BMI** | 1.15 | 0.87, 1.52 | 0.3 |
| **Age (years)** | 0.94 | 0.88, 1.00 | 0.07 |
| **Sex (female)** | 1.01 | 0.54, 1.89 | 1.0 |
| **Race/Ethnicity** |  |  |  |
| **Non-Latino Asian** | 1.43 | 0.49, 4.17 | 0.5 |
| **Non-Latino Black** | 1.38 | 0.53, 3.55 | 0.5 |
| **Non-Latino White** | 0.89 | 0.46, 1.72 | 0.7 |
| **Treated at TCH^2^** | 1.31 | 0.73, 2.37 | 0.4 |
| **Diagnosis year** | 1.02 | 0.92, 1.12 | 0.8 |
| *^1^*OR = Odds Ratio, CI = Confidence Interval  ^2^TCH = Texas Children’s Hospital | | | |

**Table S3: Hyperleukocytosis and Outcomes**

| **Characteristic** | **OR***^1^* | **95% CI***^1^* | **p-value** | **Adjustment Variables** |
| --- | --- | --- | --- | --- |
| **CNS3 Status** | 2.44 | 0.89, 6.68 | 0.08 | Z-Score BMI, Age, Sex, Diagnosis Year, Treatment Site, Cytogenetic Risk |
| **EOI MRD** | 1.3 | 0.71, 2.38 | 0.4 | Z-Score BMI, Age, Sex, Diagnosis Year, Treatment Site, Cytogenetic Risk |
|  | **HR***^1^* | **95% CI***^1^* | **p-value** | **Adjustment Variables** |
| **Overall Survival** | 1.0 | 0.29, 3.44 | 0.99 | Z-Score BMI, Age, Sex, CNS Status, Treatment Site, Diagnosis Year, Cytogenetic Risk |
| **Event Free Survival** | 0.52 | 0.19, 1.45 | 0.21 | Z-Score BMI, Age, Sex, CNS Status, Treatment Site, Diagnosis Year, Cytogenetic Risk |
| *^1^*OR, Odds Ratio. HR, Hazard Ratio, CI = Confidence Interval | | | |  |

| 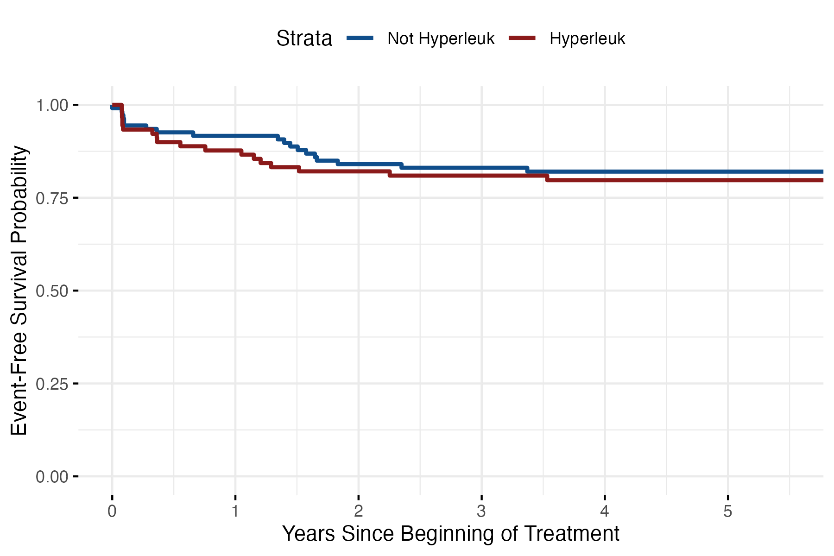  A) | 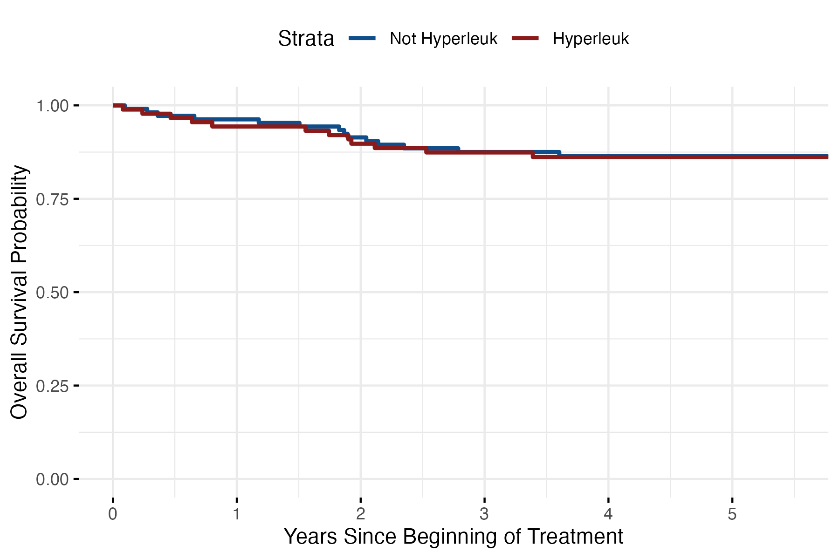  B) |
| --- | --- |
| **Figure S1.** Treatment outcomes for patients with T-ALL by hyperleukocytosis at diagnosis for A) event-free and b) overall survival. | |
